# Machine Learning-Driven Glycoproteomic Profiling Identifies Novel Diabetes-Associated Glycosylation Biomarkers

**DOI:** 10.1101/2025.02.24.25322514

**Authors:** Amr Elguoshy, Keiko Yamamoto, Yoshitoshi Hirao, Kengo Yanagita, Tomohiro Uchimoto, Takashi Kamimura, Tetsuya Takazawa, Tadashi Yamamoto

## Abstract

Glycosylation plays a critical role in protein function and disease progression, including diabetes mellitus. This study performed a comprehensive glycoproteomic analysis comparing healthy volunteers (HV) and DM samples, identifying 19,374 peptides and 2,113 proteins, of which 1,104 were glycosylated. A total of 287 distinct glycans were mapped to 3,722 glycosylated peptides, revealing significant differences in glycosylation patterns between HV and DM samples. Statistical analysis identified 29 significantly altered glycosylation sites, with 23 upregulated in DM and 6 downregulated in DM. Notably, the glycan HexNAc(2)Hex(2)Fuc(1) at position 215 of Prosaposin was significantly upregulated in DM, marking its first reported association with diabetes. Machine learning models, particularly Support Vector Machines (SVM) and Generalized Linear Models (GLM), achieved high classification accuracy (∼ 92%: 96%) in distinguishing HV and DM samples based on glycosylation features (Glycans, Glycosylated proteins, and Glycosylation sites). These findings suggest that altered glycosylation patterns may serve as potential biomarkers for diabetes-related pathophysiology and therapeutic targeting.

## Introduction

Glycosylation, a ubiquitous and intricate post-translational modification, plays a pivotal role in regulating a multitude of biological processes, including protein folding, stability, cellular signaling, and immune response^1 2^. This enzymatic process, which involves the attachment of glycans to proteins or lipids, is highly sensitive to cellular metabolic states and environmental cues. Consequently, dysregulated glycosylation patterns have been linked to the pathogenesis of chronic diseases, particularly metabolic disorders such as diabetes mellitus ^3 4 5^. For instance, disruptions in glycan biosynthesis pathways, such as impaired sialylation or fucosylation, are known to exacerbate insulin resistance and vascular dysfunction in DM ^6^

Diabetes mellitus is a chronic metabolic disorder characterized by persistent hyperglycemia resulting from defects in insulin secretion, insulin action, or both^7^. Beyond its well-documented metabolic dysregulation, DM is associated with profound changes in protein glycosylation, which contribute to disease progression and the development of complications such as nephropathy, neuropathy, retinopathy, and cardiovascular disease^6 8^. Emerging evidence suggests that glycosylation changes in DM influence key pathological processes, including chronic inflammation, insulin resistance, and endothelial dysfunction ^9 10^. As a result, specific N-glycan profiles have garnered attention as potential biomarkers for early diagnosis, prognosis, and therapeutic monitoring in DM^11 3^.

Despite significant advances in glycobiology, a comprehensive understanding of glycosylation alterations at both the protein and site-specific levels in DM remains elusive. Most studies to date have focused on global glycan profiling, leaving a critical gap in knowledge regarding the precise molecular mechanisms underlying glycosylation changes and their functional consequences in DM^3^. For example, glycosylation at specific residues can dictate protein stability, ligand binding, or subcellular localization—features critical to pathological processes in DM^12^. Addressing this gap requires advanced analytical approaches capable of capturing the complexity of glycosylation at high resolution.

In this study, we conducted a systematic and in-depth analysis of glycosylation data derived from healthy individuals and DM patients. Utilizing a combined approach of statistical analysis and machine learning, we aimed to identify dysregulated glycoproteins, glycans and glycosylation sites associated with DM^13^. This dual-level analysis provided a comprehensive view of glycosylation changes in DM.

Our analytical workflow incorporated rigorous statistical methods to identify differentially glycosylated features without imputation, followed by machine learning analyses on imputed data to evaluate the robustness and reproducibility of our findings. This integrated approach ensured the reliability of our results while offering insights into the potential impact of missing data imputation on the identification of significant glycosylation features.

Here, we present the key findings from our analysis, highlighting specific glycoproteins and glycosylation sites that are significantly associated with DM pathology. We also discuss the broader implications of these findings for the development of novel biomarkers and therapeutic targets, which could pave the way for improved management and treatment of DM.

## Materials and Methods

### 1. Sample Collection and Preparation

A total of 40 biological samples were collected, comprising 20 samples from healthy volunteers (HV) and 20 from diabetic patients (Table1). Sample collection followed standardized protocols to minimize pre-analytical variability.

**Table 1.** presents the demographic and clinical characteristics of the samples used in the study, categorized into healthy volunteers (HV) and diabetes mellitus (DM) patients. The table includes details on sample name, age, sex, HbA1c levels, and urinary protein status. **Age:** The HV and DM group includes individuals with age averages of 54.4, and 58.4 years respectively; **Sex:** Both groups comprise male and female participants.;**HbA1c Levels:** As expected, the DM group exhibits higher HbA1c values, indicative of their diabetic status.;**Urinary Protein Status:** Represented as (−), (±), and (+), indicating negative, borderline, or positive urinary protein levels, respectively.

| sample_category | age | sex | HbA1c | urinary protein |
| --- | --- | --- | --- | --- |
| HV | 51-55 | M | 5.8 | (±) |
| HV | 46-50 | M | 5.4 | (−) |
| HV | 56-60 | M | 5.5 | (−) |
| HV | 56-60 | F | 6.1 | (±) |
| HV | 51-55 | M | 5.5 | (−) |
| HV | 56-60 | M | 5.4 | (−) |
| HV | 56-60 | M | 5.6 | (−) |
| HV | 66-70 | M | 5.6 | (−) |
| HV | 56-60 | F | 5.4 | (−) |
| HV | 46-50 | M | 5.6 | (±) |
| HV | 66-70 | M | 5.5 | (±) |
| HV | 41-45 | F | 5.3 | (−) |
| HV | 61-65 | F | 6.5 | (−) |
| HV | 41-45 | M | 5.4 | (−) |
| HV | 41-45 | F | 5.3 | (−) |
| HV | 51-55 | M | 5.3 | (−) |
| HV | 56-60 | M | 5.9 | (±) |
| HV | 51-55 | F | 5.5 | (−) |
| HV | 56-60 | M | 5.3 | (−) |
| HV | 36-40 | F | 5.1 | (−) |
| DM | 41-45 | M | 9 | (−) |
| DM | 41-45 | M | 7.8 | (−) |
| DM | 26-30 | M | 5.6 | (+) |
| DM | 51-55 | M | 6.1 | (−) |
| DM | 61-66 | F | 8 | (−) |
| DM | 46-50 | F | 9.5 | (−) |
| DM | 41-45 | M | 6.7 | (±) |
| DM | 66-70 | M | 7.7 | (−) |
| DM | 56-60 | F | 8.1 | (−) |
| DM | 46-50 | F | 8.6 | (+) |
| DM | 61-65 | F | 7.1 | (−) |
| DM | 71-75 | F | 7 | (-) |
| DM | 61-65 | M | 13.4 | (-) |
| DM | 86-90 | F | 8.9 | (+) |
| DM | 66-70 | F | 8.5 | (-) |
| DM | 51-55 | F | 8.3 | (-) |
| DM | 56-60 | F | 7.4 | (-) |
| DM | 71-75 | M | 6.2 | (-) |
| DM | 71-75 | F | 7.5 | (-) |
| DM | 56-60 | M | 6.9 | (-) |

Urinary proteins, including glycoproteins, were precipitated using methanol/chloroform precipitation. Briefly, 500 µL of urine was used for each sample. Frozen urine samples were thawed at 37 °C in a water bath for 10 minutes prior to processing. An equal volume of methanol and one-quarter volume of chloroform were added to the urine sample, followed by thorough mixing for 5 minutes. The mixture was centrifuged at 19,000 × g at 25 °C for 15 minutes. The supernatant was carefully discarded without disturbing the protein-containing interface layer using a pipette. An equal volume of methanol was then added to the sample, mixed gently for 5 minutes, and centrifuged again at 19,000 × g at 25 °C for 15 minutes. The supernatant was removed, and the precipitated proteins were air-dried.

### 2. Tryptic Peptide Preparation

The precipitated proteins, including glycoproteins, were dissolved in 100 µL of 8 M urea/50 mM Tris-HCl (pH 8.0) buffer. Reduction was performed by adding 1 µL of 1 M dithiothreitol (DTT) and incubating at room temperature (RT) for 1 hour. Alkylation was carried out by adding 8 µL of 500 mM iodoacetamide (IAA) and incubating at RT for 1 hour in the dark. The alkylation reaction was quenched by adding 1 µL of 1 M DTT. The sample was then diluted eightfold with 50 mM Tris-HCl (pH 8.0). For digestion, 1 µg of trypsin (Agilent, Santa Clara, CA, USA) was added, and the sample was incubated at 37 °C for 16 hours with shaking. Digestion was terminated by acidification with 50% trifluoroacetic acid (TFA).

The digested peptides were purified using a C18 spin column (GL Science, Tokyo, Japan) according to the manufacturer’s instructions. Briefly, the C18 spin column was activated sequentially with 100% and 50% acetonitrile (ACN), followed by equilibration with 0.2% formic acid (FA) by centrifugation at 3,000 × g for 30 seconds. The sample was loaded onto the column and centrifuged at 3,000 × g for 90 seconds. Trapped peptides, including glycopeptides, were washed twice with 0.2% TFA and eluted with 95% ACN containing 5% FA. The eluted peptides were dried using a VEC-260 vacuum dryer (Iwaki, Tokyo, Japan). The dried peptides were reconstituted in 0.1% FA, and peptide concentration was measured using a NanoDrop 1000 spectrophotometer (Thermo Fisher Scientific, Bremen, Germany). Samples were stored at -80 °C until further analysis.

### 3. Mass Spectrometry Analysis

Peptide samples were analyzed using a Fusion Lumos mass spectrometer equipped with an Orbitrap detector (Thermo Fisher Scientific, Bremen, Germany) in data-dependent acquisition (DDA) mode. Peptides, including glycopeptides, were separated using a nanoflow liquid chromatography system (nLC1000, Thermo Fisher Scientific) on a trap column (2 cm × 75 µm, Acclaim PepMap 100) and an analytical column (12.5 cm × 75 µm, NTCC-360) at a flow rate of 300 nL/min. The mobile phases consisted of (A) 0.1% FA in water and (B) 0.1% FA in ACN. A total of 500 ng of peptides was injected and eluted using a linear gradient from 2% to 35% mobile phase B over 120 minutes.

The mass spectrometer was operated in positive ion mode with a scan range of 350–1800 m/z. The 20 most intense peaks with charge states of 2+, 3+ and 4+ were selected for fragmentation, with an automatic gain control (AGC) target of 4.0 × 10^5 and a maximum injection time of 50 ms. To enhance glycopeptide identification, a dual fragmentation strategy was employed, utilizing higher-energy collisional dissociation (HCD) and electron-transfer/higher-energy collisional dissociation (EThcD) for complementary fragmentation of glycopeptides^14^.

Peptides, including both deglycosylated and glycopeptides, were identified with a stringent 1% false discovery rate (FDR) at the protein level. Glycosylation sites were mapped using a combination of automated database searches and manual validation to ensure high-confidence site localization.

### 4. Database Search Parameters

MS/MS spectra were analyzed using the Byonic v3.4.0 search engine (Protein Metrics, Inc.) against the Homo sapiens SwissProt database (Version: 2018-07). The search parameters were as follows:

- Parent ion mass tolerance: 10 ppm
- Fragment ion mass tolerance: 0.02 Da
- Fixed modification: Carbamidomethylation (C)
- Variable modifications

- 309 curated mammalian N-glycans
- 6 most common O-glycans

Spectral counts of glycosylated and non-glycosylated peptides were extracted for downstream quantitative analyses.

### 4. Quantification of Glycosylation at Different Levels

Relative abundance normalization was applied to compute glycosylation levels at three hierarchical levels: site, glycan, and glycosylated protein.

- **Site-Level Glycosylation Relative Abundance:**

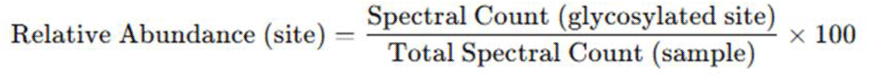
- **Glycan-Level Glycosylation Relative Abundance**

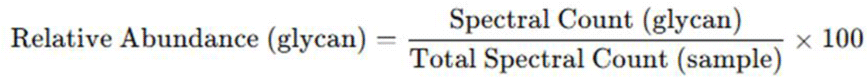
- **Glycosylated Protein-Level Glycosylation Relative Abundance**

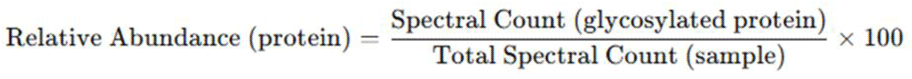

This relative abundance-based normalization accounted for variations in sample loading and MS performance, ensuring comparability across samples.

### 5. Statistical Analysis

Prior to missing data imputation, statistical tests were applied to detect significant differences in glycosylation patterns between HV and DM groups.

- Differential Glycosylation Analysis:
  ∘ Relative abundances of glycans, glycosylated proteins, and glycosylation sites were compared.
  ∘ The Wilcoxon rank-sum test was used due to non-Gaussian Relative abundances distributions.
  ∘ Fold-change analysis was performed, considering |log_2_ (fold change)| > 0.58 as biologically significant.

### 6. Missing Data Handling and Imputation

To improve dataset robustness, glycans, glycosylated proteins, and glycosylation sites detected in fewer than 10 of the 40 samples were excluded to reduce noise before applying imputation methods.

Missing values were imputed using five independent strategies to minimize bias in downstream analyses:

1. Mean imputation (substitution with group mean)
2. k-Nearest Neighbors (KNN) imputation (leveraging data similarity)
3. Random Forest imputation (preserving complex relationships)
4. Minimum imputation (assuming missing values correspond to low abundance)
5. Mixed imputation, which combines KNN for missing at random (MAR) data and minimum imputation for missing not at random (MNAR) data

Each imputation method generated a distinct dataset, and their performance was evaluated to ensure imputed values did not distort biologically meaningful patterns.

### 7. Machine Learning Analysis

Machine learning models were developed to classify HV and DM samples based on glycosylation features.

- Feature Selection:
  ∘ Boruta Algorithm was used to identify the most discriminative glycosylation features. Boruta algorithm is an extension of the Random Forest algorithm and is particularly useful for high-dimensional datasets.
- Model Training and Evaluation:
  ∘ Algorithms applied: Logistic Regression, Decision Trees, Random Forest (RF), and Support Vector Machines (SVM).
  ∘ Performance was evaluated using 10-fold cross-validation (CV) and a test dataset.
  ∘ The following metrics were calculated:
    ▪ Accuracy (overall classification performance)
    ▪ Precision and Recall (class-wise performance)
    ▪ F1-score (harmonic mean of precision and recall)
    ▪ Area Under the Receiver Operating Characteristic Curve (AUC-ROC)
- Final Model Evaluation Criteria:
  ∘ Test Performance Priority: Emphasizing test-set metrics over CV metrics for real-world applicability.
  ∘ Consistency across Metrics: Prioritizing models with balanced accuracy, precision, recall, and F1-score.
  ∘ Variance in Cross-Validation: Models with lower standard deviation (CV_SD) in performance were favored.
- Feature Importance Analysis:
  ∘ Logistic Regression coefficients and Random Forest feature importance scores were analyzed.
  ∘ Key glycosylation sites contributing to classification were identified.

### 8. Software and Computational Tools

All analyses were conducted in R (version 4.2.0). The following R packages were utilized:

∘ dplyr (data manipulation)
∘ ggplot2 (visualization)
∘ caret (machine learning modeling)
∘ missForest and DMwR (missing data imputation)
∘ pROC (ROC curve analysis)

Mass spectrometry data were processed using Byonic software, followed by statistical and machine learning analysis in R.

## Results and Discussion

### 1. Glycosylation Profiles in Healthy Volunteers (HV) and Diabetes Mellitus Samples

A comprehensive glycoproteomic analysis identified 19,374 peptides (both glycosylated and non-glycosylated), corresponding to 2,113 proteins across all samples. Among these, 287 distinct glycans were mapped to 3,722 glycosylated peptides, covering 9,194 unique glycosylation sites and 1,104 glycosylated proteins. Specifically, 282 N-glycans and 5 O-glycans were identified. This comprehensive dataset underscores the complexity of glycosylation in both physiological and pathological states, consistent with prior studies highlighting the diversity of glycan structures in human proteomes15. The predominance of N-glycans aligns with their established role in stabilizing protein conformations and mediating cellular interactions, processes frequently disrupted in metabolic diseases like DM^2^.

Notably, aberrant glycosylation in DM has been linked to immune dysregulation and impaired cell signaling^6^. Our findings reinforce this paradigm, revealing diabetes-specific glycoforms that may contribute to disease progression. For instance, the identification of 5 O-glycans, though fewer in number, suggests potential roles in modulating extracellular matrix interactions or inflammatory pathways, as observed in other chronic inflammatory conditions^16^.

### 2. Dysregulated Glycans, Glycoproteins and Glycosylation Sites in DM

Comparative statistical analysis revealed significant differences in glycosylation patterns between HV and DM cohorts (p < 0.05). Among 1,700 shared glycosylation sites, 29 exhibited altered abundances (23 upregulated in DM; 6 downregulated in DM) ((Figures 1a–1c).), while 22 of 576 common glycoproteins were dysregulated (16 upregulated in DM; 6 downregulated in DM) (Figures 2a–2c) (Tables-2).

**Figure 1(a–c):**
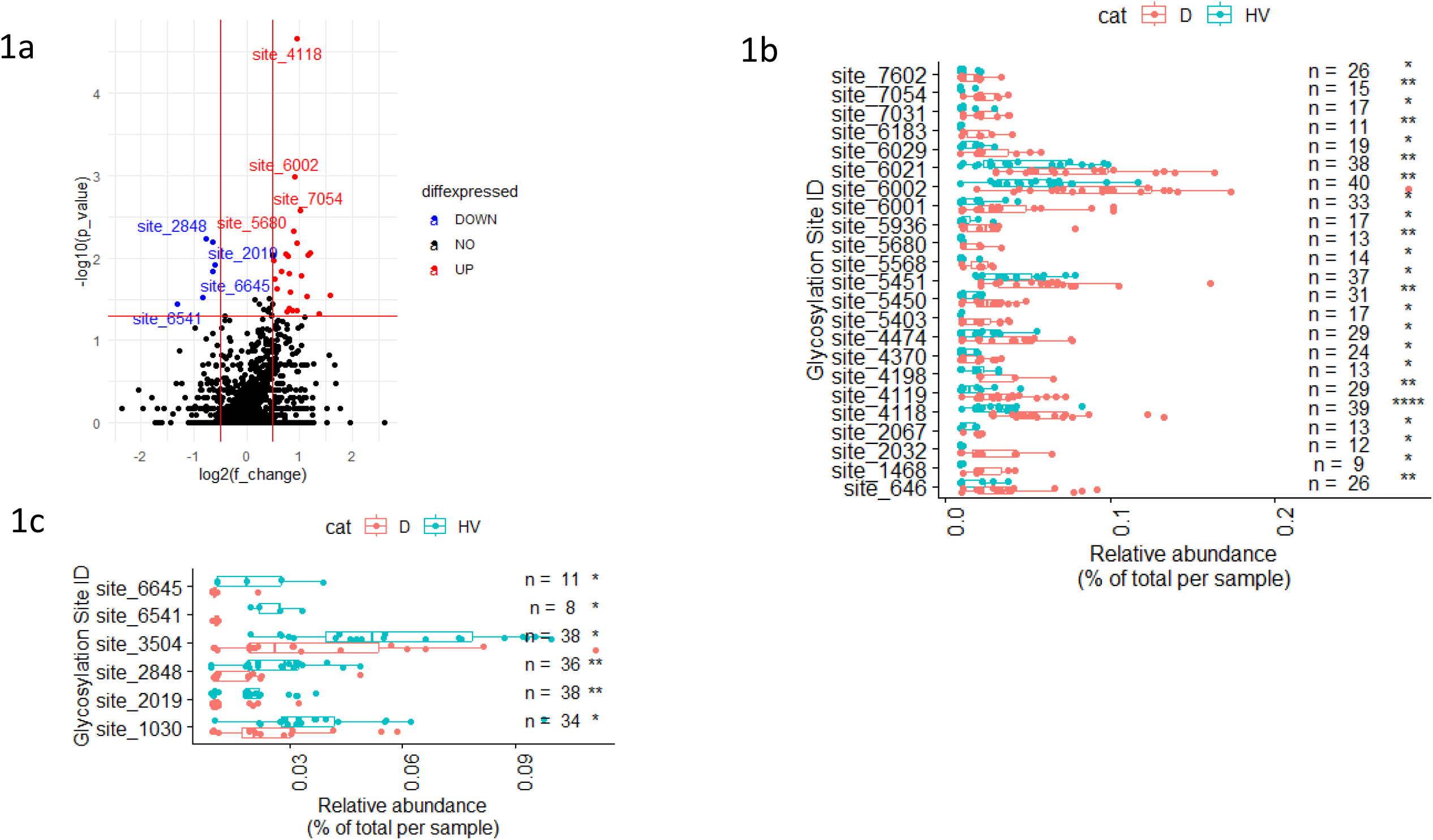
Comparative analysis of glycosylation sites between healthy volunteers (HV) and diabetes mellitus (DM) samples. **1a**: Volcano plot displaying significantly dysregulated glycosylation sites in DM compared to HV (p < 0.05). **1b**: Boxplots highlighting upregulated glycans at specific glycosylation sites in DM. **1c**: Boxplots showing downregulated glycans at specific glycosylation sites in DM.

**Figure 2(a–c):**
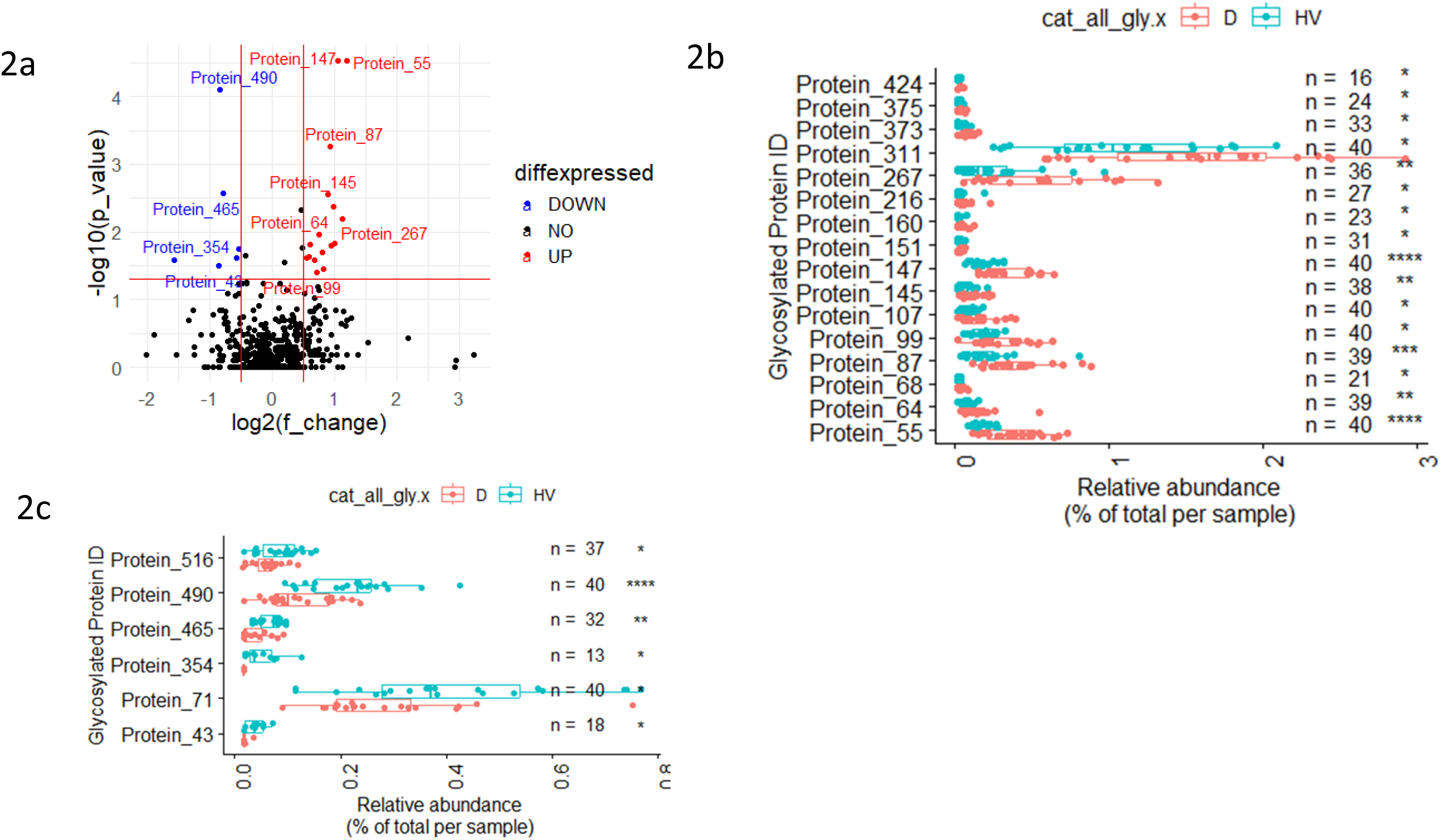
Comparative analysis of glycosylated proteins between healthy volunteers (HV) and diabetes mellitus (DM) samples. **2a**: Volcano plot displaying significantly dysregulated glycosylated proteins in DM compared to HV (p < 0.05). **2b**: Boxplots highlighting upregulated glycosylated proteins in DM. **2c**: Boxplots showing downregulated glycosylated proteins in DM.

Key findings include the upregulation of the glycan HexNAc(2)Hex(2)Fuc(1) at position 215 of the Prosaposin precursor protein (site_4118) in DM (p < 0.0001) (Figure 1a – 1b), representing the first report of this glycan’s association with this specific site in DM. This glycan, previously detected in unrelated contexts such as healthy human urine^17^, Gaucher spleen tissue ^18^, and GM1 gangliosidosis urine ^19^ at undefined positions, has never been linked to diabetes with the glycosylation at position 215.

Four of the five identified sphingolipid activator proteins (SAPs), namely SAP-A, SAP-B, SAP-C, and SAP-D, originate from a common precursor protein (Prosaposin) through proteolytic processing. The SAP-precursor is encoded by a gene located on chromosome 10, which comprises 15 exons and 14 introns. Importantly, glycosylation at position 215 is critical for SAP-B function in lysosomal sphingolipid degradation, as its loss reduces protein stability and activity, leading to lysosomal storage disorders. This novel finding emphasizes its potential as a biomarker for diabetes-related lysosomal dysfunction ^20^.

HexNAc(5)Hex(6)Fuc(1)NeuAc(1)NeuGc(1) at position 78 in Prostaglandin-H2 D-isomerase (site_7054) (p < 0.01) (Figure 1a, 1b) (Table 2), which was previously identified in prostate cancer urine; and HexNAc(4)Hex(5)NeuAc(2) at position 115 in AMBP precursor protein (site_6021) (p < 0.01) (Figure 1a, 1b). These observations suggest that glycosylation changes in DM may overlap with other pathological states, but their specific roles in DM-related processes, such as inflammation and oxidative stress, warrant further investigation.

**Table 2.** an overview of the identified glycosylation sites, glycosylated proteins, and glycans associated with diabetes mellitus (DM). The data includes unique glycan feature IDs (used in figures) and their corresponding names, and status in DM (UP: increased, DOWN: decreased).

| type | ID | Name | status in DM |
| --- | --- | --- | --- |
| Glycosylation site | site_4118 | HexNAc(2)Hex(2)Fuc(1); Prosaposin ;215 | UP |
| Glycosylation site | site_6002 | HexNAc(4)Hex(5)NeuAc(2); Prothrombin ;143 | UP |
| Glycosylation site | site_7054 | HexNAc(5)Hex(6)Fuc(1)NeuAc(1)NeuGc(1); Prostaglandin-H2 D-isomerase;78 | UP |
| Glycosylation site | site_5680 | HexNAc(4)Hex(5)Fuc(2)NeuAc(1); Zinc-alpha-2-glycoprotein ;128 | UP |
| Glycosylation site | site_2848 | HexNAc(1)Hex(1)NeuAc(2); Protein YIPF3 ;346 | DOWN |
| Glycosylation site | site_2019 | HexNAc(1)Hex(1)NeuAc(1); Phosphoinositide-3-kinase-interacting protein 1 ;38 | DOWN |
| Glycosylation site | site_4119 | HexNAc(2)Hex(2)Fuc(1); Prosaposin ;332 | UP |
| Glycosylation site | site_6183 | HexNAc(4)Hex(6)Fuc(1)NeuAc(1); Protein AMBP ;115 | UP |
| Glycosylation site | site_6021 | HexNAc(4)Hex(5)NeuAc(2); Protein AMBP ;115 | UP |
| Glycosylation site | site_646 | HexNAc(1)Hex(1); Alpha-2-HS-glycoprotein ;346 | UP |
| Glycosylation site | site_5450 | HexNAc(4)Hex(5)Fuc(1)NeuAc(1); Prostaglandin-H2 D-isomerase;51 | UP |
| Glycosylation site | site_2067 | HexNAc(1)Hex(1)NeuAc(1); Protein YIPF3 ;346 | UP |
| Glycosylation site | site_1030 | HexNAc(1)Hex(1); Phosphoinositide-3-kinase-interacting protein 1 ;38 | DOWN |
| Glycosylation site | site_5451 | HexNAc(4)Hex(5)Fuc(1)NeuAc(1); Prostaglandin-H2 D-isomerase;78 | UP |
| Glycosylation site | site_3504 | HexNAc(2)Fuc(1); Non-secretory ribonuclease ;119 | DOWN |
| Glycosylation site | site_7031 | HexNAc(5)Hex(6)Fuc(1)NeuAc(1); Prostaglandin-H2 D-isomerase;78 | UP |
| Glycosylation site | site_6001 | HexNAc(4)Hex(5)NeuAc(2); Prothrombin ;121 | UP |
| Glycosylation site | site_4370 | HexNAc(2)Hex(5); Immunoglobulin heavy constant alpha 1 ;340 | UP |
| Glycosylation site | site_7602 | HexNAc(6)Hex(3)Fuc(1)NeuAc(1); Polymeric immunoglobulin receptor ;469 | UP |
| Glycosylation site | site_5568 | HexNAc(4)Hex(5)Fuc(1)NeuAc(2); Glutaminyl-peptide cyclotransferase ;49 | UP |
| Glycosylation site | site_2032 | HexNAc(1)Hex(1)NeuAc(1); N-acetylmuramoyl-L-alanine amidase ;168 | UP |
| Glycosylation site | site_5403 | HexNAc(4)Hex(5)Fuc(1); Prostaglandin-H2 D-isomerase ;78 | UP |
| Glycosylation site | site_6645 | HexNAc(5)Hex(4)NeuAc(1); Prostaglandin-H2 D-isomerase ;78 | DOWN |
| Glycosylation site | site_6541 | HexNAc(5)Hex(4)Fuc(1)NeuAc(2); Kininogen-1 ;294 | DOWN |
| Glycosylation site | site_4474 | HexNAc(2)Hex(6); Lysosomal alpha-glucosidase ;470 | UP |
| Glycosylation site | site_5936 | HexNAc(4)Hex(5)NeuAc(1); Zinc-alpha-2-glycoprotein ;128 | UP |
| Glycosylation site | site_6029 | HexNAc(4)Hex(5)NeuAc(2); Hemopexin ;453 | UP |
| Glycosylation site | site_4198 | HexNAc(2)Hex(3); Lysosomal acid phosphatase ;133 | UP |
| Glycosylation site | site_1468 | HexNAc(1)Hex(1)NeuAc(1); Immunoglobulin kappa light chain ;114 | UP |
| Glycosylated protein | Protein_55 | >sp\|P00734\|THRB_HUMAN Prothrombin OS=Homo sapiens OX=9606 GN=F2 PE=1 SV=2 | UP |
| Glycosylated protein | Protein_147 | >sp\|P07602\|SAP_HUMAN Prosaposin OS=Homo sapiens OX=9606 GN=PSAP PE=1 SV=2 | UP |
| Glycosylated protein | Protein_490 | >sp\|Q96FE7\|P3IP1_HUMAN Phosphoinositide-3-kinase-interacting protein 1 OS=Homo sapiens OX=9606 GN=PIK3IP1 PE=1 SV=2 | DOWN |
| Glycosylated protein | Protein_87 | >sp\|P01876\|IGHA1_HUMAN Immunoglobulin heavy constant alpha 1 OS=Homo sapiens OX=9606 GN=IGHA1 PE=1 SV=2 | UP |
| Glycosylated protein | Protein_465 | >sp\|Q8NBJ4\|GOLM1_HUMAN Golgi membrane protein 1 OS=Homo sapiens OX=9606 GN=GOLM1 PE=1 SV=1 | DOWN |
| Glycosylated protein | Protein_145 | >sp\|P07339\|CATD_HUMAN Cathepsin D OS=Homo sapiens OX=9606 GN=CTSD PE=1 SV=1 | UP |
| Glycosylated protein | Protein_64 | >sp\|P01009\|A1AT_HUMAN Alpha-1-antitrypsin OS=Homo sapiens OX=9606 GN=SERPINA1 PE=1 SV=3 | UP |
| Glycosylated protein | Protein_267 | >sp\|P25311\|ZA2G_HUMAN Zinc-alpha-2-glycoprotein OS=Homo sapiens OX=9606 GN=AZGP1 PE=1 SV=2 | UP |
| Glycosylated protein | Protein_373 | >sp\|Q07075\|AMPE_HUMAN Glutamyl aminopeptidase OS=Homo sapiens OX=9606 GN=ENPEP PE=1 SV=3 | UP |
| Glycosylated protein | Protein_68 | >sp\|P01024\|CO3_HUMAN Complement C3 OS=Homo sapiens OX=9606 GN=C3 PE=1 SV=2 | UP |
| Glycosylated protein | Protein_311 | >sp\|P41222\|PTGDS_HUMAN Prostaglandin-H2 D-isomerase OS=Homo sapiens OX=9606 GN=PTGDS PE=1 SV=1 | UP |
| Glycosylated protein | Protein_160 | >sp\|P08473\|NEP_HUMAN Neprilysin OS=Homo sapiens OX=9606 GN=MME PE=1 SV=2 | UP |
| Glycosylated protein | Protein_71 | >sp\|P01042\|KNG1_HUMAN Kininogen-1 OS=Homo sapiens OX=9606 GN=KNG1 PE=1 SV=2 | DOWN |
| Glycosylated protein | Protein_424 | >sp\|Q5IJ48\|CRUM2_HUMAN Protein crumbs homolog 2 OS=Homo sapiens OX=9606 GN=CRB2 PE=1 SV=2 | UP |
| Glycosylated protein | Protein_216 | >sp\|P15586\|GNS_HUMAN N-acetylglucosamine-6-sulfatase OS=Homo sapiens OX=9606 GN=GNS PE=1 SV=3 | UP |
| Glycosylated protein | Protein_151 | >sp\|P07858\|CATB_HUMAN Cathepsin B OS=Homo sapiens OX=9606 GN=CTSB PE=1 SV=3 | UP |
| Glycosylated protein | Protein_516 | >sp\|Q9GZM5\|YIPF3_HUMAN Protein YIPF3 OS=Homo sapiens OX=9606 GN=YIPF3 PE=1 SV=1 | DOWN |
| Glycosylated protein | Protein_354 | >sp\|P80370\|DLK1_HUMAN Protein delta homolog 1 OS=Homo sapiens OX=9606 GN=DLK1 PE=1 SV=3 | DOWN |
| Glycosylated protein | Protein_375 | >sp\|Q08174\|PCDH1_HUMAN Protocadherin-1 OS=Homo sapiens OX=9606 GN=PCDH1 PE=1 SV=2 | UP |
| Glycosylated protein | Protein_43 | >sp\|O75891\|AL1L1_HUMAN Cytosolic 10-formyltetrahydrofolate dehydrogenase OS=Homo sapiens OX=9606 GN=ALDH1L1 PE=1 SV=2 | DOWN |
| Glycosylated protein | Protein_107 | >sp\|P02790\|HEMO_HUMAN Hemopexin OS=Homo sapiens OX=9606 GN=HPX PE=1 SV=2 | UP |
| Glycosylated protein | Protein_99 | >sp\|P02760\|AMBP_HUMAN Protein AMBP OS=Homo sapiens OX=9606 GN=AMBP PE=1 SV=1 | UP |
| Glycan | Glycan_80 | HexNAc(4)Hex(5)NeuAc(1) | UP |
| Glycan | Glycan_83 | HexNAc(4)Hex(5)NeuGc(1) | UP |
| Glycan | Glycan_87 | HexNAc(4)Hex(6)Fuc(1)NeuAc(1) | UP |
| Glycan | Glycan_98 | HexNAc(5)Hex(3)Fuc(1) | UP |

Conversely, downregulated glycans in DM included: HexNAc(1)Hex(1)NeuAc(1) O-glycan at position Thr-38 in Phosphoinositide-3-kinase-interacting protein precursor (site_2019) (p < 0.01) (Figure 1a, 1c) (Table 2), previously reported as O-linked in prostate cancer urine ^21^, and HexNAc(1)Hex(1)NeuAc(2) O-glycan at position Thr-346 in YIPF3 precursor protein (site_2848) (p < 0.01) (Figure 1a, 1c), also identified in urine from a single study^22^. The observed downregulation may suggest an impairment of these glycosylation pathways in diabetes, with potential consequences for protein function and disease progression.

Among dysregulated glycoproteins, Prosaposin (GN=PSAP) (Protein_147) and Prothrombin (GN=F2) (Protein_55) were the most significantly upregulated in DM (p < 0.0001) (Figure 2a, 2b) (Table 2), while Phosphoinositide-3-kinase-interacting protein (GN=PIK3IP1) (Protein_490) (p < 0.0001) and Golgi membrane protein (GN=GOLM1) (Protein_465) (p < 0.01) were the most significantly downregulated ((Figure 2a, 2c)). Reduced glycosylation here may impair these protein signaling, a pathway central to insulin sensitivity.

Among 268 shared glycans, the most significantly upregulated in DM was HexNAc(4)Hex(5)NeuAc(1) (Glycan_80) (p < 0.0001), followed by HexNAc(4)Hex(5)NeuGc(1) (Glycan_83) and HexNAc(5)Hex(3)Fuc(1) (Glycan_98) (p < 0.01) (Figure 3a) (Table 2). These dysregulated glycosylation patterns and proteins are strongly linked to processes such as inflammation, oxidative stress, and glucose metabolism, reinforcing their relevance as potential biomarkers or therapeutic targets in DM. The identification of specific glycans and their precise roles in DM pathophysiology provides a foundation for further studies to explore their diagnostic and therapeutic potential.

**Figure 3a:**
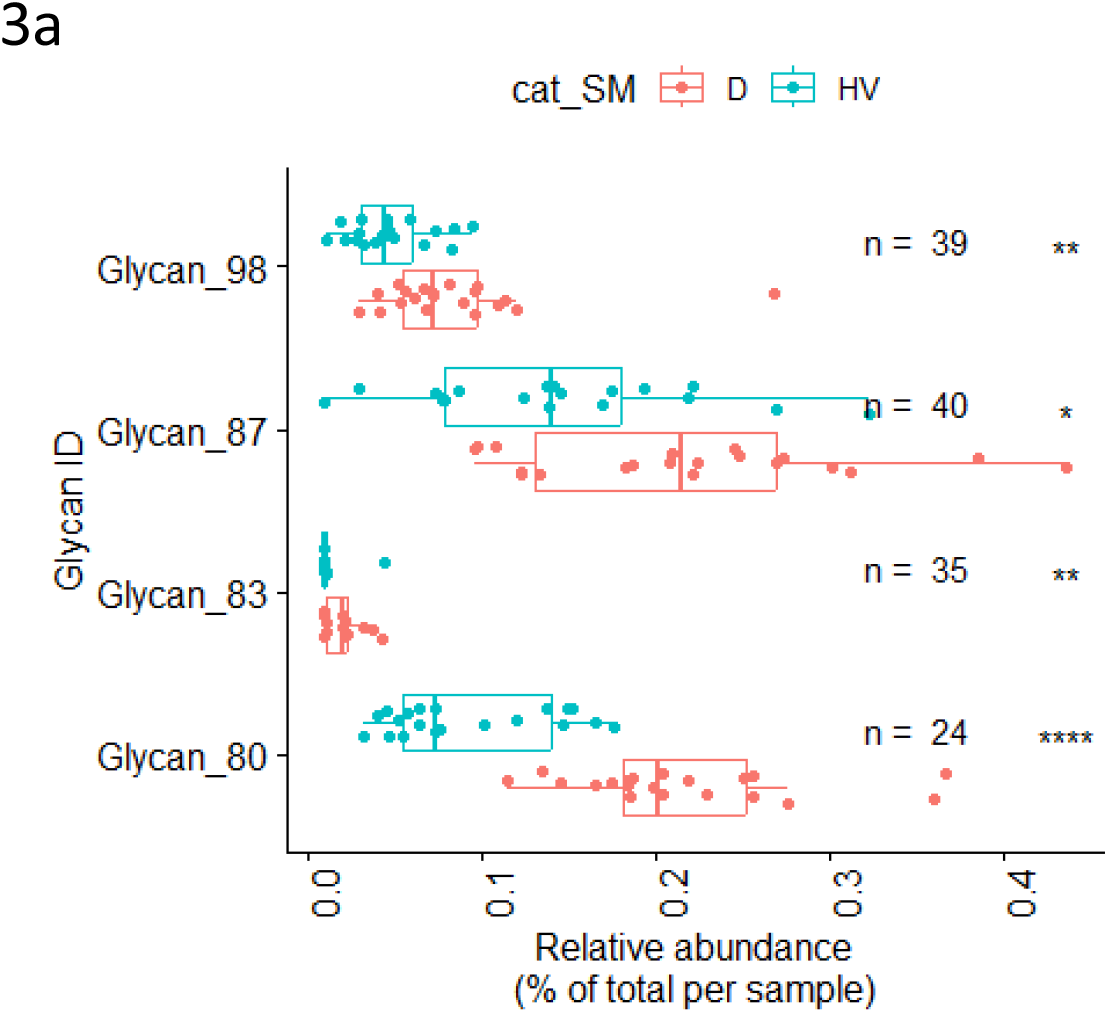
Comparative analysis of the identified glycans between healthy volunteers (HV) and diabetes mellitus (DM) samples. **3a**: Boxplots highlighting upregulated glycans in DM compared to HV.

### 3. Data Filtration and Imputation

Data filtration retained glycans, glycosylated proteins and glycosylation sites detected in at least 10 out of 40 samples, resulting in 197 distinct glycans, 223 glycosylated proteins, and 450 glycosylation sites. This represented 73%, 38%, and 26% of the shared glycans, proteins, and sites, respectively, highlighting the challenge of analyzing low-abundance glycoproteins in clinical contexts.

Missing data analysis revealed proportions of 34.2%, 41.7%, and 49.7% for glycans, proteins, and sites, respectively.

To address missing data, five imputation methods were applied, generating distinct datasets for downstream analyses for each level (Glycosylation sites, Glycosylated proteins, Glycans). The impact of each method on data distribution was evaluated to determine the most suitable approach for our highly right-skewed dataset.

Both mean imputation (Figures 4a, 5a, 6a) and MissForest imputation (Figures 4c, 5c, 6c) resulted in noticeable distortions of the data distribution, with a tendency to overestimate missing values. The similarity between the results of MissForest and mean imputation suggests that missing values may be randomly distributed and do not strongly depend on other variables. This indicates that the imputation model lacks sufficient features or variability to make accurate predictions.

**Figures 4(a–e):**
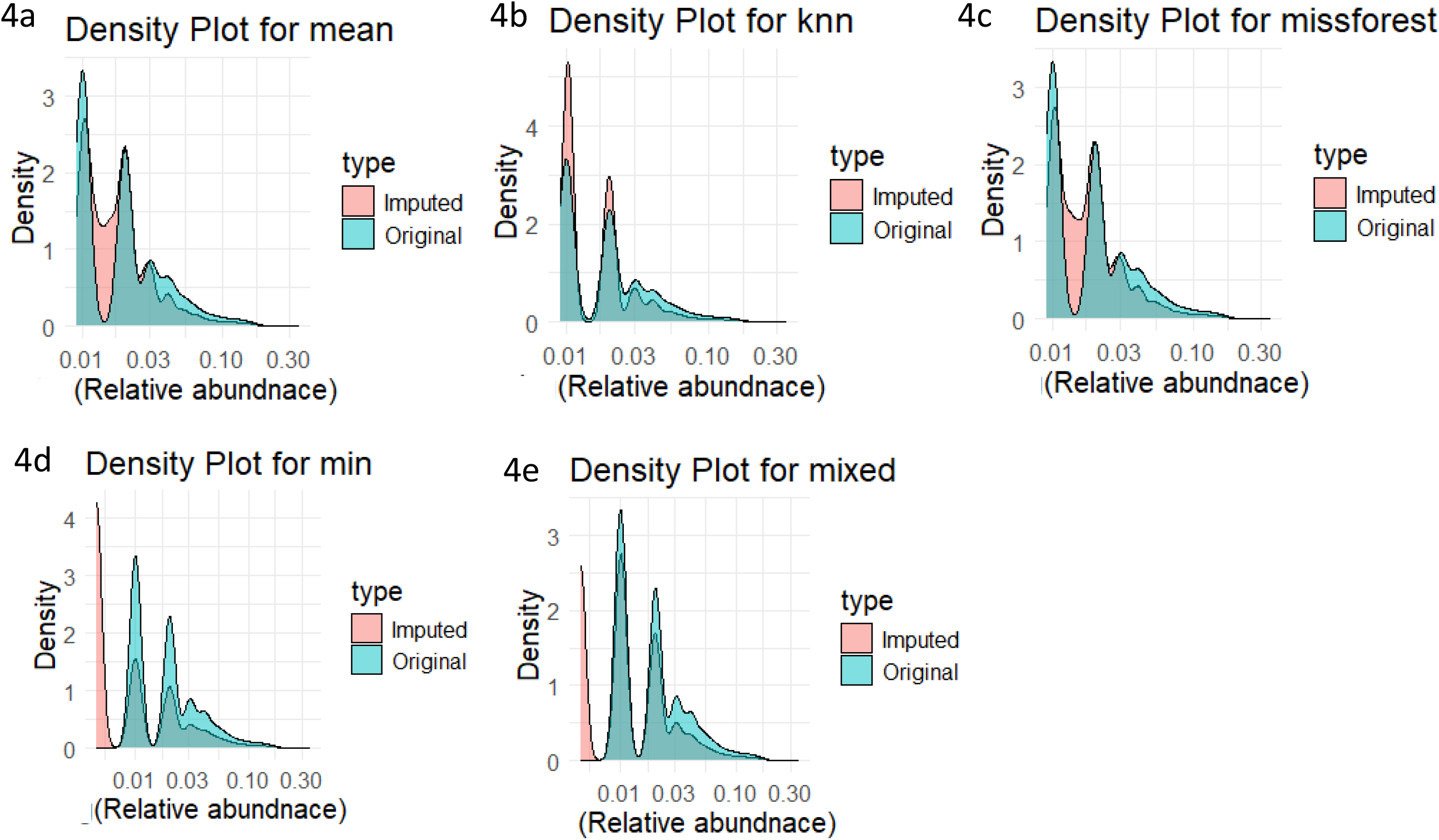
Comparison of different imputation methods for glycosylation site data. **4a**: Mean imputation effects on glycan data distribution. **4b**: K-nearest neighbors (KNN) imputation. **4c**: MissForest imputation. **4d**: Minimum imputation. **4e**: Mixed imputation.

**Figures 5(a–e):**
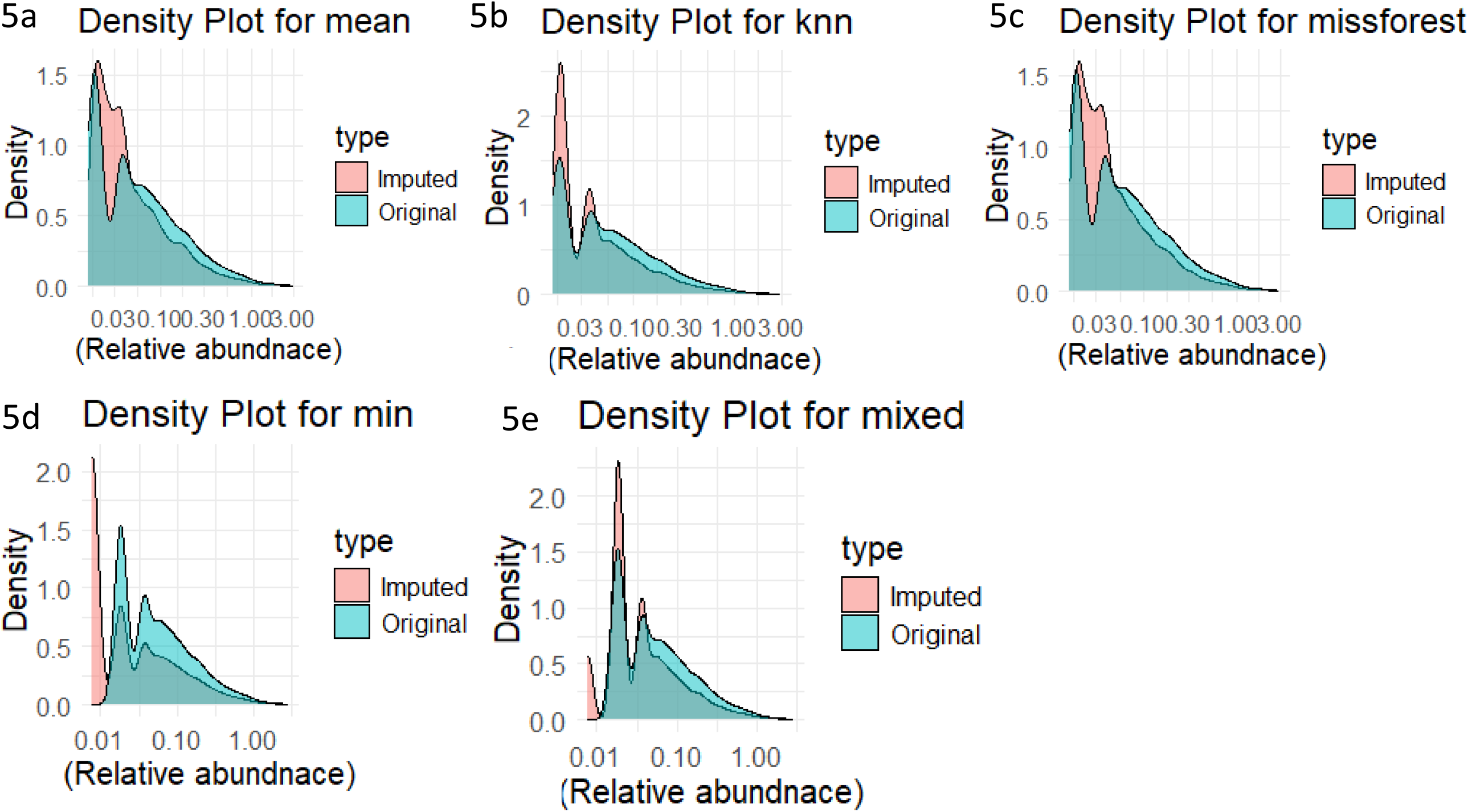
Comparison of different imputation methods for glycosylated protein data. **5a**: Mean imputation effects on glycan data distribution. **5b**: K-nearest neighbors (KNN) imputation. **5c**: MissForest imputation. **5d**: Minimum imputation. **5e**: Mixed imputation.

**Figures 6(a–e):**
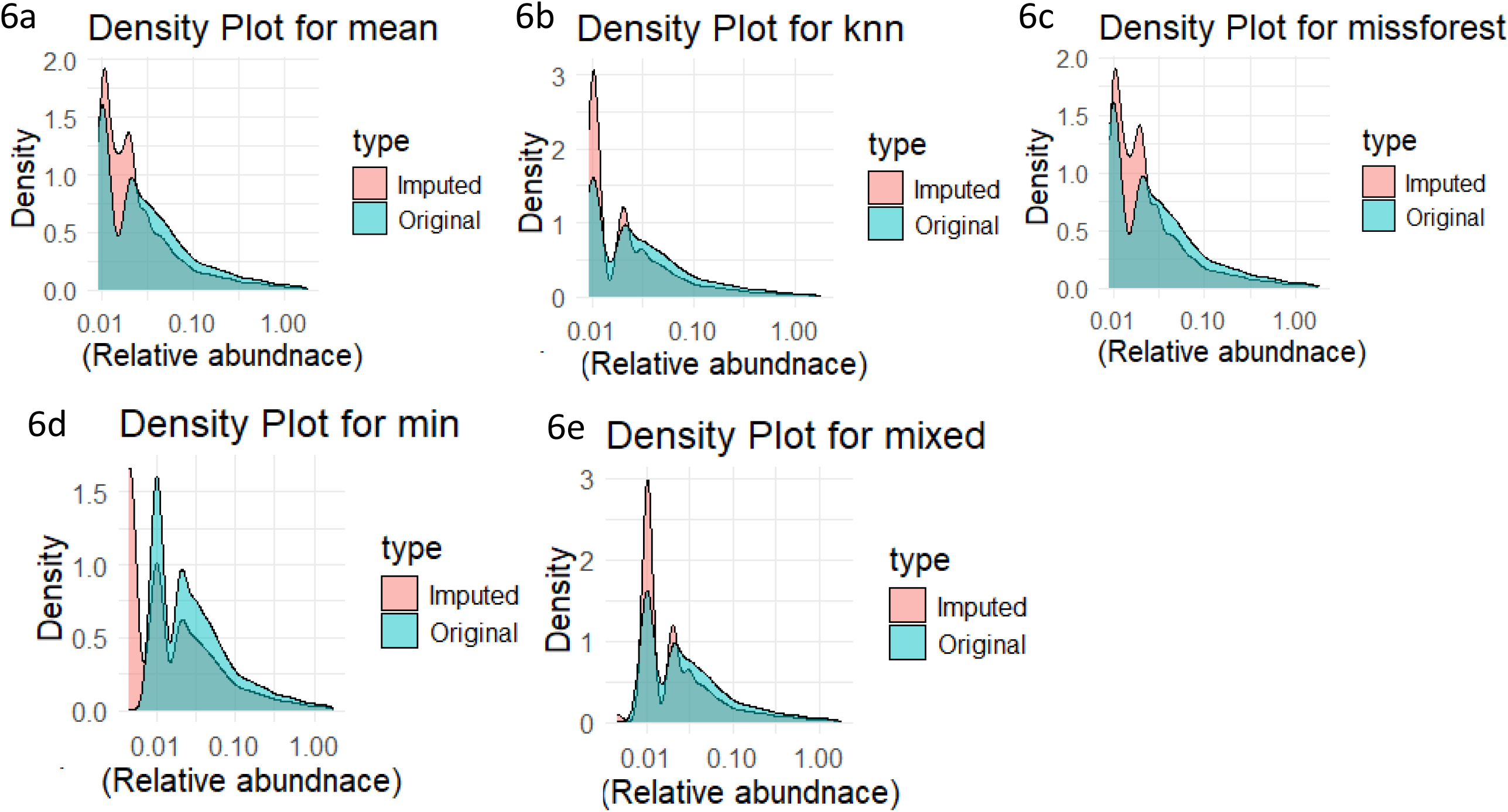
Comparison of different imputation methods for glycan data. **6a**: Mean imputation effects on glycan data distribution. **6b**: K-nearest neighbors (KNN) imputation. **6c**: MissForest imputation. **6d**: Minimum imputation. **6e**: Mixed imputation.

In contrast, K-nearest neighbors (KNN) imputation (4b, 5b, 6b) produced imputed values that were better distributed and more closely aligned with the original data structure. This method demonstrated greater adaptability to the inherent variability in the dataset.

Minimum imputation (4d, 5d, 6d) preserved the overall distribution but introduced a pronounced peak at the lower extreme of the data range, likely leading to an underestimation of missing values. Similarly, mixed imputation (4e, 5e, 6e), which combines KNN for missing at random (MAR) data and minimum imputation for missing not at random (MNAR) data, maintained the data distribution while introducing a small peak at the lower end. Although this approach may also underestimate missing values, it aligns well with the intrinsic right-skewed nature of the dataset, where most real values are small.

Given the highly right-skewed nature of the data, methods incorporating minimum and mixed imputation are expected to yield the most reliable results. However, the final assessment of imputation performance should be based on the evaluation of machine learning models trained on these imputed datasets, as model performance will ultimately determine the most suitable imputation strategy for our data.

## 4. Machine Learning Analysis

### Glycosylation Sites

Feature selection using Boruta identified between 17 (missForest) and 31 (KNN) significant glycosylation sites out of 450 features. Four sites (“gly_2019,” “gly_4118,” “gly_5450,” and “gly_3504”) were consistently selected across all methods. Support Vector Machine (SVM) models trained on minimum-imputed data achieved optimal classification performance, with an average cross-validation accuracy of 96%, precision of 94%, recall of 100%, specificity of 92%, F1-score of 97%, and AUC-ROC of 96%. The test set performance was similarly robust across all metrics (Figure 7a, 7b).

**Figures 7(a, b):**
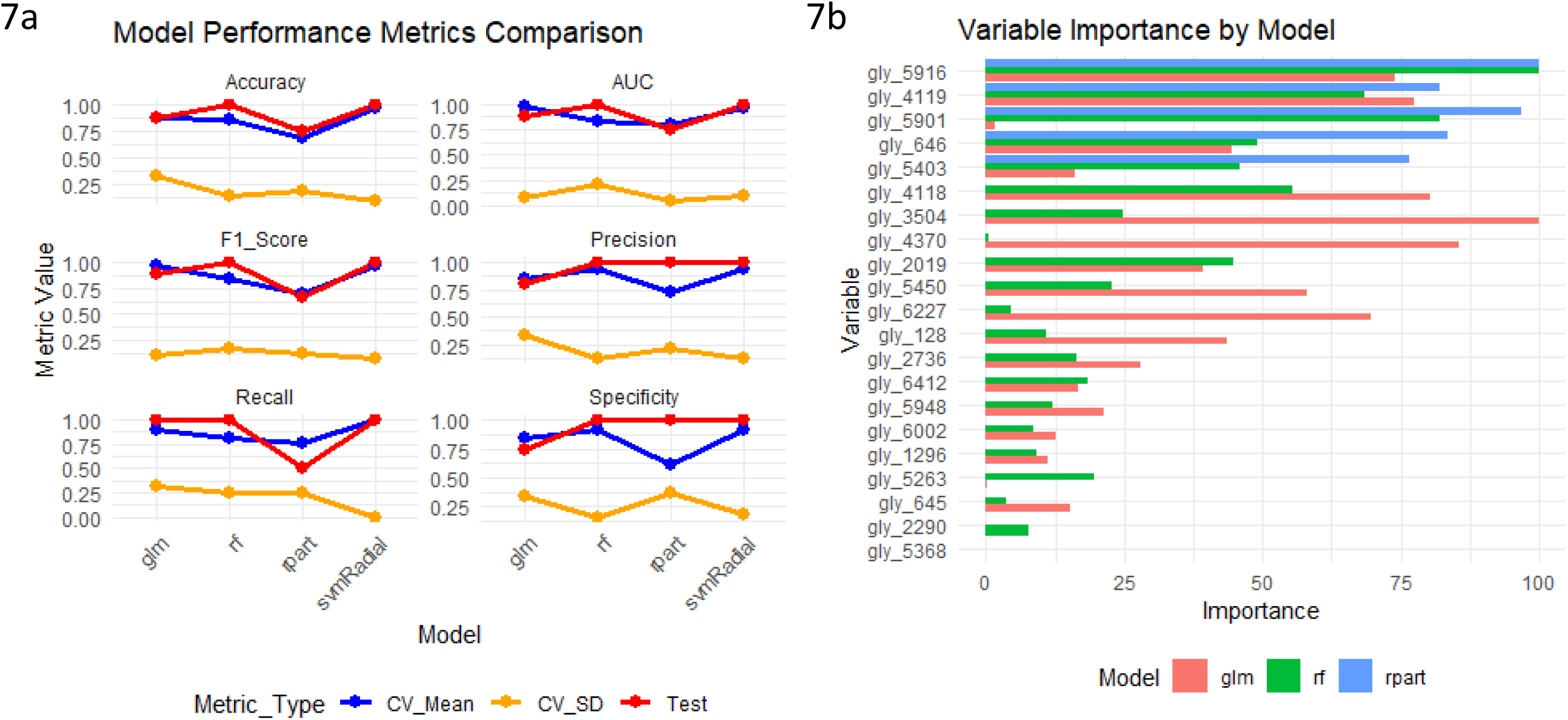
Machine Learning Classification Performance and Variable Importance for Glycosylation Site Data Imputed by Minimum Imputation. **7a**: Feature importance analysis, identifying the glycosylation sites that contributed most to the classification. **7b**: Performance metrics of the classification model, including accuracy, precision, recall, specificity, F1-score, and AUC-ROC for both cross-validated (CV) and test data. Since minimum imputation produced the most accurate and reliable results, performance metrics for other imputation methods are not shown.

#### Glycosylated Proteins

Feature selection retained 15 to 21 glycoproteins, depending on the imputation method, with nine proteins consistently identified. SVM models trained on mixed-imputed data achieved the highest performance, yielding a cross-validation accuracy of 92%, precision of 93%, and AUC-ROC of 92%. Test set metrics reached 100% accuracy, precision, recall, and AUC (Figure 8a, 8b).

**Figures 8(a, b):**
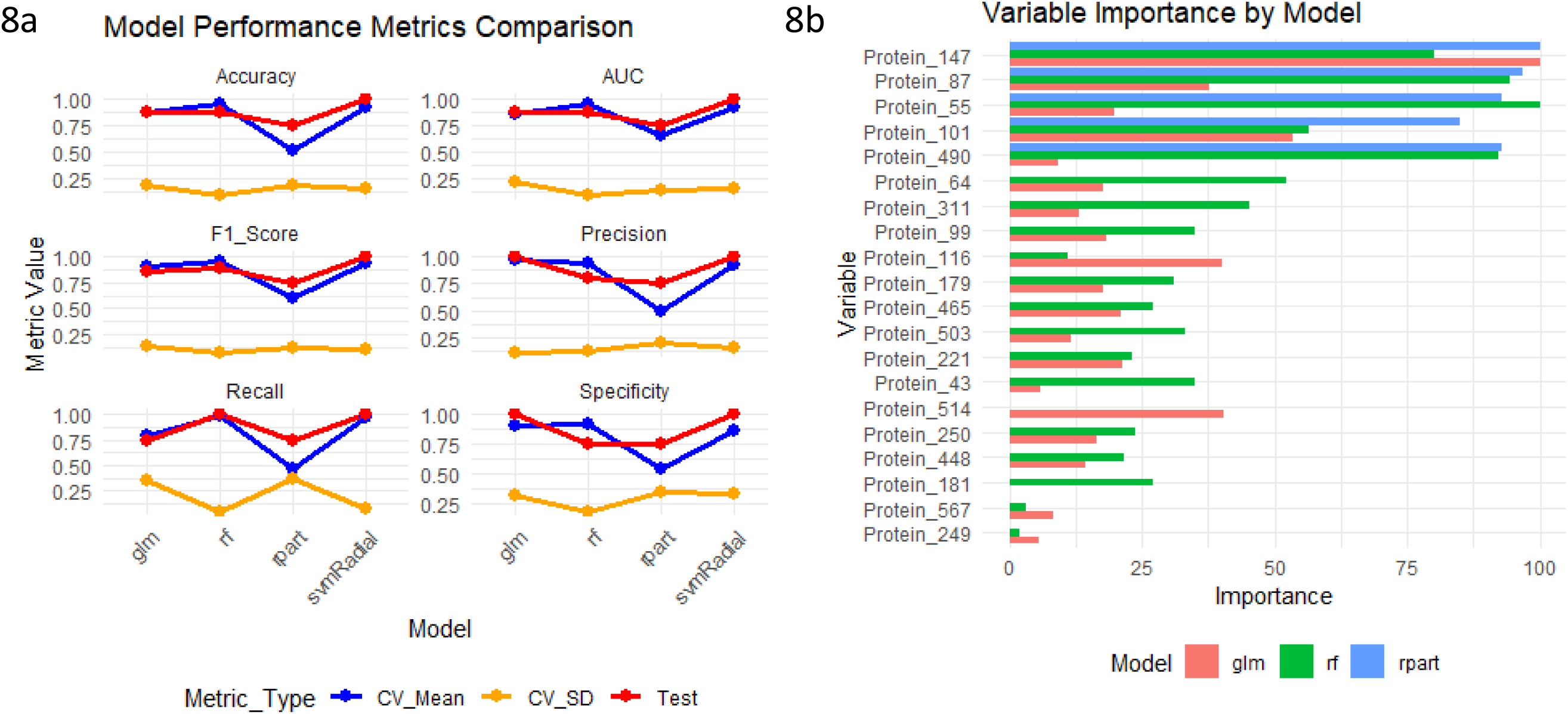
Machine Learning Classification Performance and Variable Importance for Glycoprotein Data Imputed by mixed Imputation. **8a**: Feature importance analysis, identifying the glycosylated proteins that contributed most to the classification. **8b**: Performance metrics of the classification model, including accuracy, precision, recall, specificity, F1-score, and AUC-ROC for both cross-validated (CV) and test data. Since mixed imputation produced the most accurate and reliable results, performance metrics for other imputation methods are not shown.

#### Glycans

Feature selection retained 9 to 12 features depending on the imputation method, with four glycans (“Glycan_18”, “Glycan_80”, “Glycan_98”, “Glycan_254”) consistently identified. Generalized linear models (GLMs) trained on KNN-imputed data achieved superior classification performance, with cross-validation accuracy of 94%, precision of 100%, and AUC-ROC of 95%. Test set performance metrics were similarly high, reinforcing the robustness of the approach (Figure 9a, 9b).

**Figures 9(a, b):**
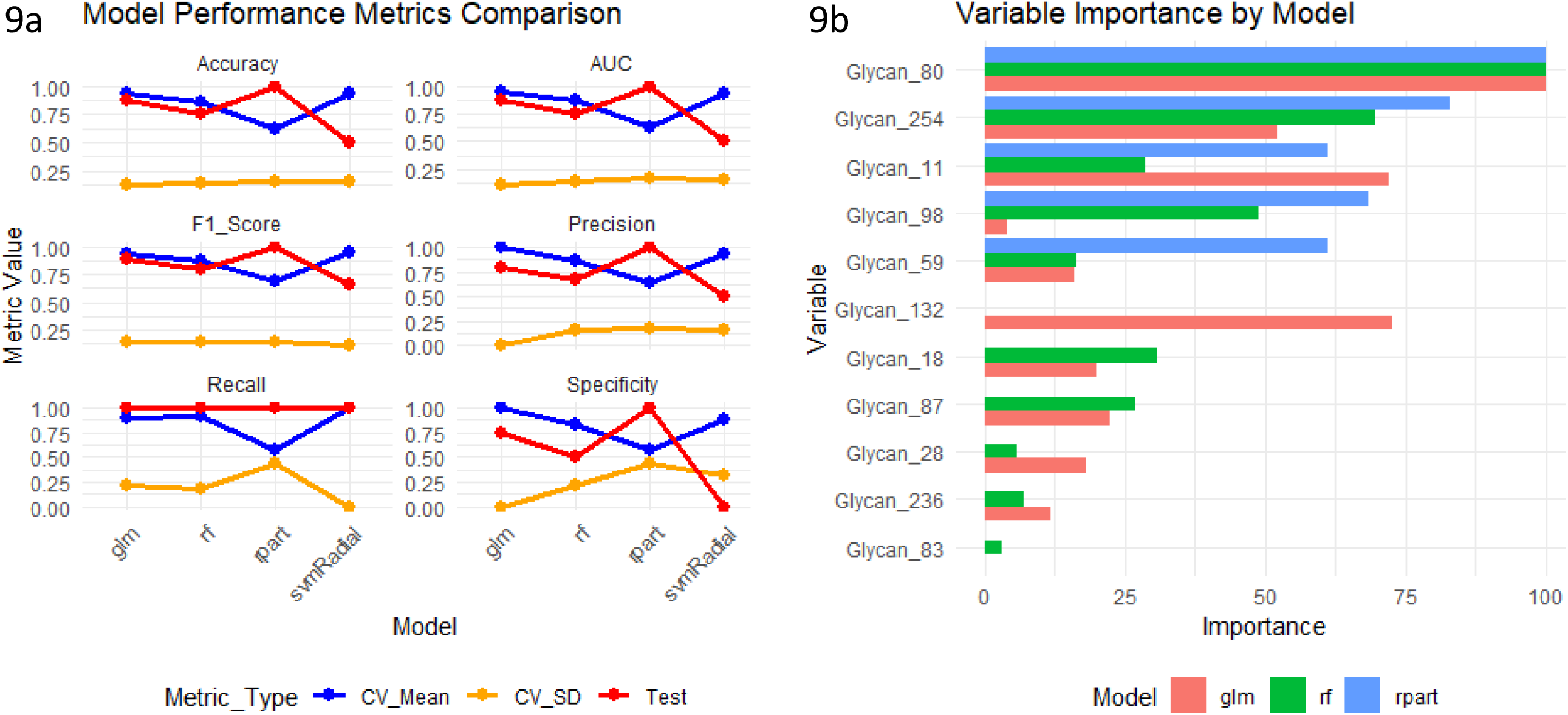
Machine Learning Classification Performance and Variable Importance for Glycan Data Imputed by K-Nearest Neighbor (KNN) Imputation. **8a**: Feature importance analysis, identifying the glycans that contributed most to the classification. **8b**: Performance metrics of the classification model, including accuracy, precision, recall, specificity, F1-score, and AUC-ROC for both cross-validated (CV) and test data. Since KNN imputation produced the most accurate and reliable results, performance metrics for other imputation methods are not shown.

Both SVM and GLM demonstrated superior performance with KNN and minimum imputation strategies, providing reliable tools for distinguishing HV and DM samples based on glycosylation signatures.

The differential performance of SVM and GLM in glycosylation data analysis is inherently linked to the complexity, structure, and statistical properties of the data. SVM excels in modeling non-linear and high-dimensional relationships prevalent in glycosylation site and glycoprotein data, whereas GLMs are more suited for compositional glycan data with linear additive effects. The integration of both approaches provides a comprehensive framework for identifying glycosylation-based biomarkers and understanding disease-specific glycosylation alterations.

The overlap between statistically significant features and machine learning priorities (e.g., Prosaposin N215) validates their biological relevance. This dual approach mirrors recent advances in integrative glycoproteomics, where hybrid frameworks enhance biomarker discovery. Dysregulated glycosylation features highlighted by these methods offer valuable insights into DM pathology and hold potential as biomarkers or therapeutic targets.

### 5. Novel Site-Specific Glycosylation in DM

In this study, we identified distinct glycosylation patterns at specific amino acid residues in proteins from individuals with Diabetes Mellitus that were absent in healthy controls. Specifically, the Asn-831 and Ser-833 residues in the Fibrinogen alpha chain were found to be systematically glycosylated exclusively in DM patients. The Asn-831 site was modified with a HexNAc(1)Fuc(1) glycan, while the Ser-833 site was modified with a HexNAc(1)Hex(1)NeuAc(1) glycan. Additionally, the Thr-231 residue in Roundabout homolog 4 (ROBO4) was observed to be systematically glycosylated exclusively in DM patients, with an O-linked glycan structure of HexNAc(1)Hex(1)NeuAc(2).

These findings represent the first reported instance of glycosylation at these specific residues in the context of DM. Notably, these glycosylation sites have not been previously documented in the GlyConnect database, underscoring their novelty and potential significance in the pathophysiology of DM. Further investigation into the functional implications of these glycosylation events may provide insights into the molecular mechanisms underlying DM and its associated complications.

## Conclusion

This study provides a detailed glycoproteomic landscape of diabetes mellitus, identifying significant alterations in glycosylation patterns. The discovery of novel glycosylation sites and their association with DM highlights the potential of glycosylation as a biomarker for disease progression. The application of statistical and machine learning methods further strengthens the robustness of these findings, demonstrating their utility in distinguishing DM from healthy states. These results pave the way for future functional investigations and biomarker validation in clinical settings.

## Data Availability

All data produced in the present study are available upon reasonable request to the authors

## Institutional Review Board Statement

This study was conducted in accordance with the Declaration of Helsinki and was approved by the ethics committees of Shinrakuen Hospital (No. H290005) and Niigata University (No. 2021-0025).

## Informed Consent Statement

Urine samples used in this study were collected from left-over specimens of healthy subjects (volunteers) after laboratory tests for their health check at Shinrakuen Hospital and were anonymized with link information unavailable to the investigators. The ethics committees approved the collection and use of these samples by obtaining informed consent in the form of opt-out.

## Acknowledgments

COI: BBC is the Collaborative Research Laboratory of Tosoh Corporation.

